# Establishing causal relationships between sleep and adiposity traits using Mendelian randomisation

**DOI:** 10.1101/2022.07.08.22277418

**Authors:** Bryony L Hayes, Marina Vabistsevits, Richard M Martin, Deborah A Lawlor, Rebecca C Richmond, Timothy Robinson

## Abstract

**Objective:** To systematically evaluate the direction of any potential causal effect between sleep and adiposity traits.

**Methods:** Two-sample Mendelian randomization (MR) was used to assess the association of genetically predicted sleep traits on adiposity and vice versa. Using data from UK Biobank and 23andme, the sleep traits explored were morning-preference (chronotype) (N=697,828), insomnia (N=1,331,010), sleep duration (N=446, 118), napping (N=452,633) and daytime-sleepiness (N=452,071). Using data from the GIANT and EGG consortia, the adiposity traits explored were adult BMI, hip circumference (HC), waist circumference (WC), waist-to-hip ratio (WHR) (N=322,154) and child-BMI (N=35,668).

**Results:** We found evidence that insomnia symptoms increased mean WC, BMI and WHR (difference in means WC=0.39 SD (95% CI=0.13, 0.64), BMI=0.47 SD (0.22, 0.73) and WHR=0.34 SD (0.16, 0.52)). Napping increased mean WHR (0.23 SD (0.08, 0.39). Higher HC, WC, and adult-BMI increased odds of daytime-sleepiness (HC=0.02 SD (0.01, 0.04), WC=0.04 SD (0.01, 0.06) and BMI 0.02 SD (0.00, 0.04), respectively). We also found that higher mean child-BMI resulted in lower odds of napping (−0.01 SD (0.02, 0.00).

**Conclusions:** The effects of insomnia on adiposity, and adiposity on daytime-sleepiness, suggest that poor sleep and weight gain may contribute to a feedback loop that could be detrimental to overall health.

## Introduction

Poor sleep is common, with up to 67% of UK adults reporting disturbed sleep, 26 – 36% experiencing insomnia and 23% sleeping for < 5 hrs per night[1]. Sleep traits, such as chronotype (i.e. morning- or evening-preference), insomnia and sleep duration, have previously been studied in relation to both being overweight and obesity. Sleep disorders and obesity have been linked to almost every aspect of health, from mental health[2–4] to overall physical health [4–9]. Therefore, establishing the extent to which they relate to each other is important for identifying modifiable targets for interventions that could have beneficial effects on healthy sleep and weight and hence other health outcomes.

Conventional multivariable regression analyses show reported evening preference, insomnia, short- and long-sleep duration to associate with increased odds of obesity (Body Mass Index (BMI) ≥30 kg/m^2^)[10][11]. However, it is difficult to determine whether these associations are causal or explained by residual confounding or reverse causality. These studies have predominantly explored whether sleep has an impact on adiposity, with few investigating whether there is a reverse relationship – a potential effect of adiposity on sleep traits.

Mendelian randomisation (MR) is a causal inference approach that utilizes germline genetic variants associated with potentially modifiable risk factors as instruments to estimate causal effects on outcomes. MR is less vulnerable to biases incurred by conventional observational analyses, such as reverse causation and confounding, though there are a set of assumptions that can produce biased estimates when violated [12–14].

We have identified three existing MR studies that have explored potential causal effects between adiposity and sleep traits [15–17] (**Table 1**). Together these suggest that higher adult BMI potentially increases daytime napping and sleepiness and morning preference, greater waist circumference and waist-to-hip ratio increase daytime napping, longer sleep duration reduces child BMI, and more frequent napping may increase waist circumference and waist-to-hip-ratio. None of these systematically explore a range of sleep traits with a range of adiposity traits within the same study, making it difficult to establish potential bidirectional effects from these separate studies, and most did not undertake sensitivity analyses to explore bias due to assumption violations.

**Table 1.** Summary of previously published two-sample MR studies for sleep and adiposity traits.

| Study | Exposure | Outcome | Sample size (outcome) | Sensitivity analyses | Beta (95% CI) | Pval |
| --- | --- | --- | --- | --- | --- | --- |
| Dashti et al 2021 [15] | Sleep Duration | BMI | 339,224 | No | 0.0393 (−0.055, 0.134) | 4.20E-01 |
|  | Daytime Napping | BMI | 339,224 |  | 0.211 (0.022, 0.40) | 3.00E-02 |
|  | Morning preference | BMI | 339,224 |  | −0.012 (−0.053, 0.030) | 5.90E-01 |
|  | Daytime Sleepiness | BMI | 339,224 |  | 0.199 (−0.332, 0.731) | 4.60E-01 |
|  | Insomnia | BMI | 339,224 |  | 0.36 (0.262, 0.458) | 1.25E-12 |
|  | BMI | Sleep Duration | 446, 118 |  | 0.014 (−0.042, 0.071) | 6.30E-01 |
|  | BMI | Daytime Napping | 452,633 |  | 0.027 (0.001, 0.052) | 4.00E-02 |
|  | BMI | Morning preference | 697,828 |  | 0.060 (0.002, 0.118) | 4.60E-02 |
|  | BMI | Daytime Sleepiness | 452,071 |  | 0.018 (0.008, 0.028) | 4.00E-04 |
|  | BMI | Insomnia | 1,331,010 |  | 0.01 (−0.010, 0.030) | 4.20E-01 |
| Dashti et al 2021 [16] | Daytime Napping | Waist Circumference | 232,101 | Limited | 0.282 (0.11, 0.453) | 1.00E-03 |
|  | Insomnia | Waist-to-hip ratio | 210,082 |  | 0.29 (0.172, 0.408) | 1.58E-07 |
|  | Sleep Duration | Obesity (clinically determined) | 16,033 |  | 0.995 (0.987, 1.003) | 2.40E-01 |
|  | Waist Circumference | Daytime Napping | 452,633 |  | 0.03 (−0.007, 0.067) | 1.10E-01 |
|  | Waist-to-hip ratio | Insomnia | 1,331,010 |  | −0.03 (−0.050, −0.010) | 7.16E-07 |
|  | Waist-to-hip ratio (adj. BMI) | Sleep Duration | 446, 118 |  | −0.14 (−0.19, −0.08) | 5.03E-06 |
| Wang et al 2019 [17] | Sleep Duration | Child BMI | 35,668 | Limited | −0.29 (−0.54, −0.04) | 2.20E-02 |

Our aim was to systematically evaluate the potential causal direction of effect between sleep and adiposity traits.

## Methods

We used two-sample MR analyses in which the associations of the germline genetic instrumental variants with both the exposure (sample 1) and the outcome (sample 2) were derived from two independent (i.e. non-overlapping) samples. Sleep traits explored in this study were: morning-preference, insomnia, sleep duration, napping during the day and daytime-sleepiness. Adiposity traits explored in this study included adult body mass index (adult-BMI), childhood body mass index (child-BMI), waist circumference (WC), hip circumference (HC), and waist-to-hip ratio (WHR).

For further information regarding study design see **Fig 1**.

**Figure 1.**
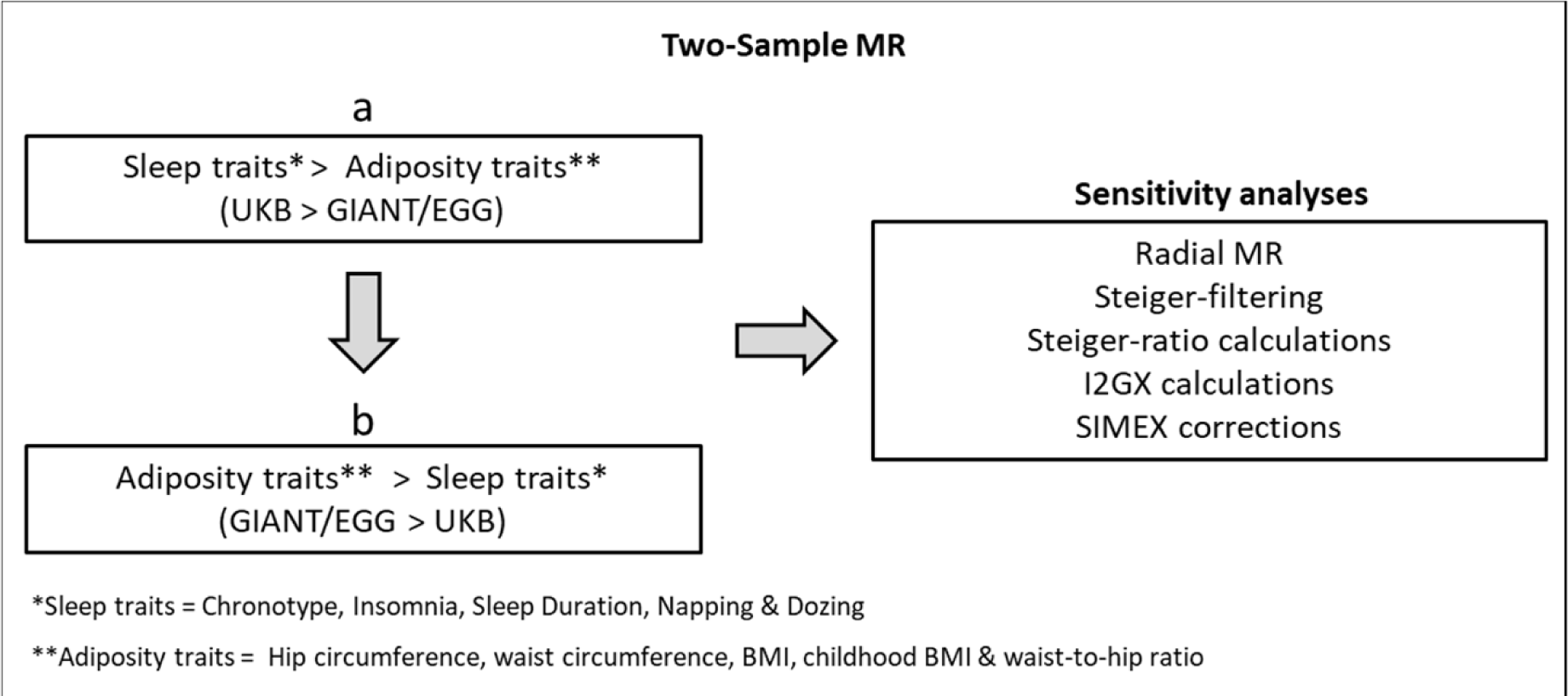
Study design. Two-sample MR is used in the main analyses to explore the relationship between sleep and adiposity, and sensitivity analyses are used to assess robustness of these associations.

### Genetic instruments for sleep traits

Genetic instruments for sleep duration, napping and daytime dozing traits used in two-sample MR were generated from GWAS conducted in UK Biobank (UKB)[18,19], and instruments for morning-preference and insomnia were generated from GWAS conducted in a meta-analysis of UKB and 23andme. For this, a linear mixed model association method was used to account for relatedness and population stratification, with BOLT-LMM software (v2.3)[20]. At baseline, participants completed a touchscreen questionnaire, which included questions about their sleep behaviours. Details of these questions are available in supplementary methods (**A**).

We identified genetic instruments for chronotype from UKB/23andme summary data (N = 697,828) [24], from which 351 independent SNPs (defined as R^2^ < 0.001) reached genome-wide significance (P<5×10^−08^). For insomnia, we identified 246 independent SNPs that reached genome-wide significance from UKB/23andme summary data (N = 1,331,010) [25]. For sleep duration (N = 446, 118)[26], napping (N = 452,633)[16], and daytime-sleepiness (N = 452,071)[27], genetic instruments were from UKB summary data, for which 78, 93 and 38 independent SNPs that achieved genome-wide significance were identified, respectively. **Supplementary table 1** provides summary statistics of the IVs used to instrument each trait.

### Genetic instruments for adiposity traits

Genetic instruments for all adult adiposity traits used in two-sample MR were generated from the Genetic Investigation of ANthropometric Traits (GIANT) consortia, a meta-analysis of ∼59 studies from across the UK and Europe[21], with those for child BMI generated from the Early Growth Genetics (EGG) consortia, a meta-analysis of ∼20 studies from across the UK and Europe[22]. BMI was calculated from weight (kg) divided by the square of height in metres (m2). An adult is classified as overweight if their BMI is 25.0 – 29.9, and obese if their BMI is >30. Measures of hip- and waist circumference were both taken in centimetres, and waist-to-hip ratio was calculated from waist circumference divided by hip circumference.

We identified genetic instruments for HC, WC, adult-BMI and WHR from GIANT consortium summary data (N = 322,154) [21,23], for which 52, 41, 68 and 29 independent (defined as R^2^ < 0.001) single nucleotide polymorphisms (SNPs) reached genome-wide significance (P<5×10^−08^) respectively. Genetic instruments for child-BMI were obtained from EGG Consortium summary data (N = 35,668) [22], for which 6 SNPs reached genome-wide significance. **Supplementary table 1** provides summary statistics of the IVs used to instrument each trait.

### Statistical Analyses

The two-sample MR approach uses genome-wide significant IVs to obtain estimates for the causal effect of risk factors on our chosen outcomes. For the univariable MR analyses in this study, the TwoSampleMR R package was used to combine and harmonize genetic summary data for each of our sleep exposure traits to determine the causal effect on adiposity, and subsequently for each of our adiposity traits to determine the causal effect on sleep. For all main analyses, an inverse variance weighted (IVW) approach was used, whereby an estimate of the causal effect is obtained from the slope of a regression line through the weighted IV-mean exposure vs IV-mean outcome associations, with the line constrained to have an intercept of zero.

#### Sensitivity Analyses and Limiting Assumption Violation

Three key assumptions must be fulfilled to ensure the validity of an MR study for making causal inference: i) the relevance assumption, that genetic IVs are statistically robustly associated with the exposure of interest in the population to which inference is made; ii) the independence assumption, that there is no confounding between the genetic IVs and outcome; and iii) the exclusion restriction assumption, that genetic IVs only influence an outcome through the exposure of interest[28].

We explored instrument strength with the F-statistics of the association between the IVs and each exposure[29,30]. Population substructure can confound genetic instrument-outcome associations and therefore, it was minimised by restricting analyses to European ancestry participants and using GWAS data that had adjusted for principal components reflecting different ancestral sub-populations. To explore the potential for unbalanced horizontal pleiotropy we conducted sensitivity analyses using MR-Egger[31], weighted median[32] and weighted mode[33] MR, and also assessed between-SNP heterogeneity using Cochran’s Q and leave one out analyses[13,34]. I^2^ statistics were used to estimate the proportion of the variance between IV estimates that is due to heterogeneity[35]. Weighted and unweighted I^2^_GX_ statistics were calculated to provide an indicator for the expected relative bias of the MR-Egger causal estimate[36], and SIMEX corrections were conducted to extrapolate bias-adjusted inference where necessary[37]. To identify IVs with the largest contribution towards heterogeneity, radial-MR was conducted (alpha = 0.05/nSNP) [38]. To identify instrumental SNPs more strongly associated with the outcome of interest than the exposure, Steiger-filtering was conducted [39]. Following both radial-MR and Steiger-filtering, MR was then repeated with any outliers removed to assess their impact.

Sample overlap between UKB and GIANT/EGG is negligible, therefore analyses between these consortia should not violate the independence assumption in two-sample MR.

MR analyses used the R package “TwoSampleMR” (version 0.5.6)[26], R version 4.0.4.

## Results

We observed an increase in WC (1 SD = 12.5cm), adult-BMI (5.1kg/m^2^) and WHR (1 SD = 0.07) for each category increase in insomnia symptoms (from never/rarely, to sometimes, to usually) (beta=0.39 SD, 95% CI=0.13, 0.64, beta=0.47 SD, 95% CI=0.22, 0.73 and beta=0.34 SD, 95% CI=0.16, 0.52 respectively) (**Fig 2**). For each hour increase in sleep duration, we observed a decrease in child-BMI (SD=4.7kg/m^2^) (beta=-0.93 SD, 95% CI=-1.74, -0.11) (**Fig 3**). For each category increase in napping, we observed an increase in WC and WHR (beta=0.28 SD, 95% CI=0.09, 0.46 and beta=0.23 SD, 95% CI=0.08, 0.39 respectively) (**Fig 4**). There was little evidence for effects of sleep traits on adiposity traits (**Figs 2 - 6**). For all analyses, results from MR-Egger, weighted median and weighted mode methods were similar to IVW estimates (**Supp Table 3**).

**Figure 2.**
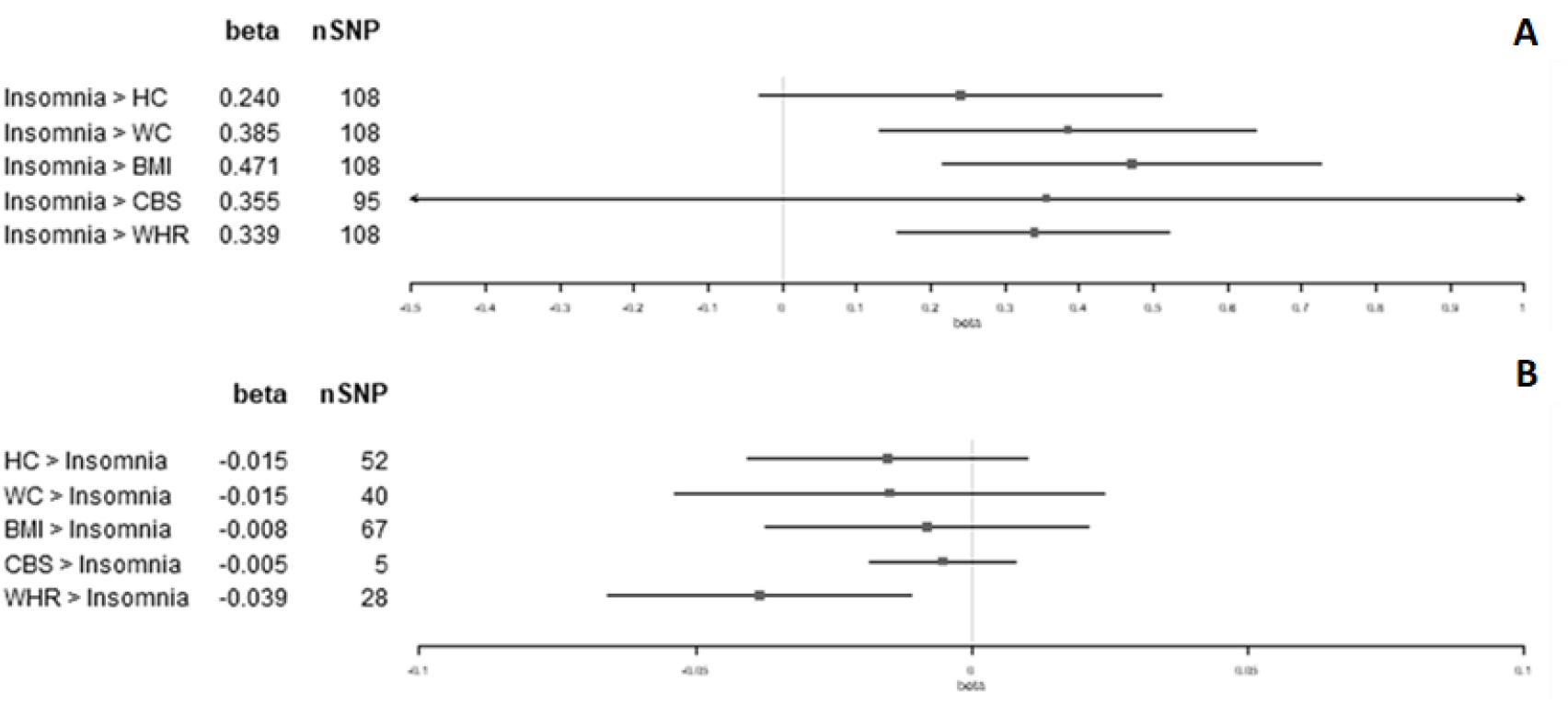
Two-sample bidirectional MR. Forest plot of A) insomnia trait effects on adiposity and B) Adiposity trait effects on insomnia. All results presented are IVW.

**Figure 3.**
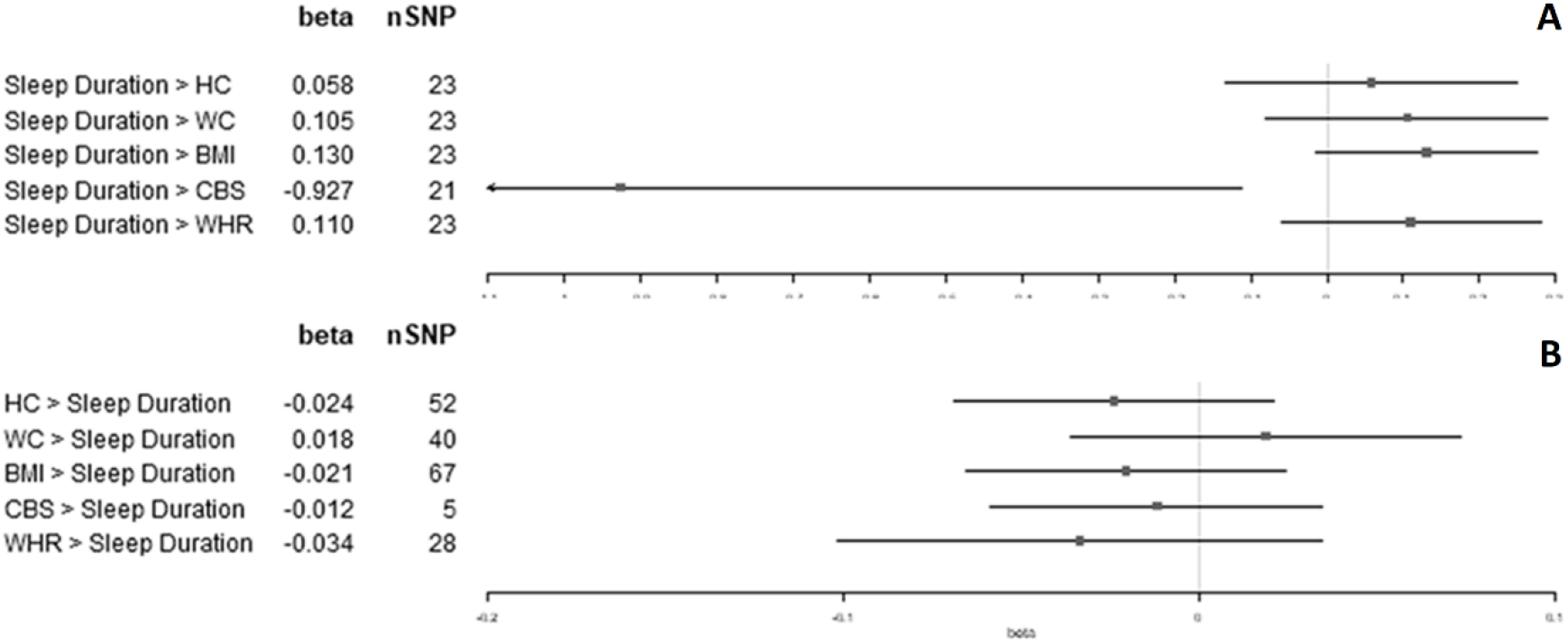
Two-sample bidirectional MR. Forest plot of A) sleep duration trait effects on adiposity and B) Adiposity trait effects on sleep duration. All results presented are IVW.

**Figure 4.**
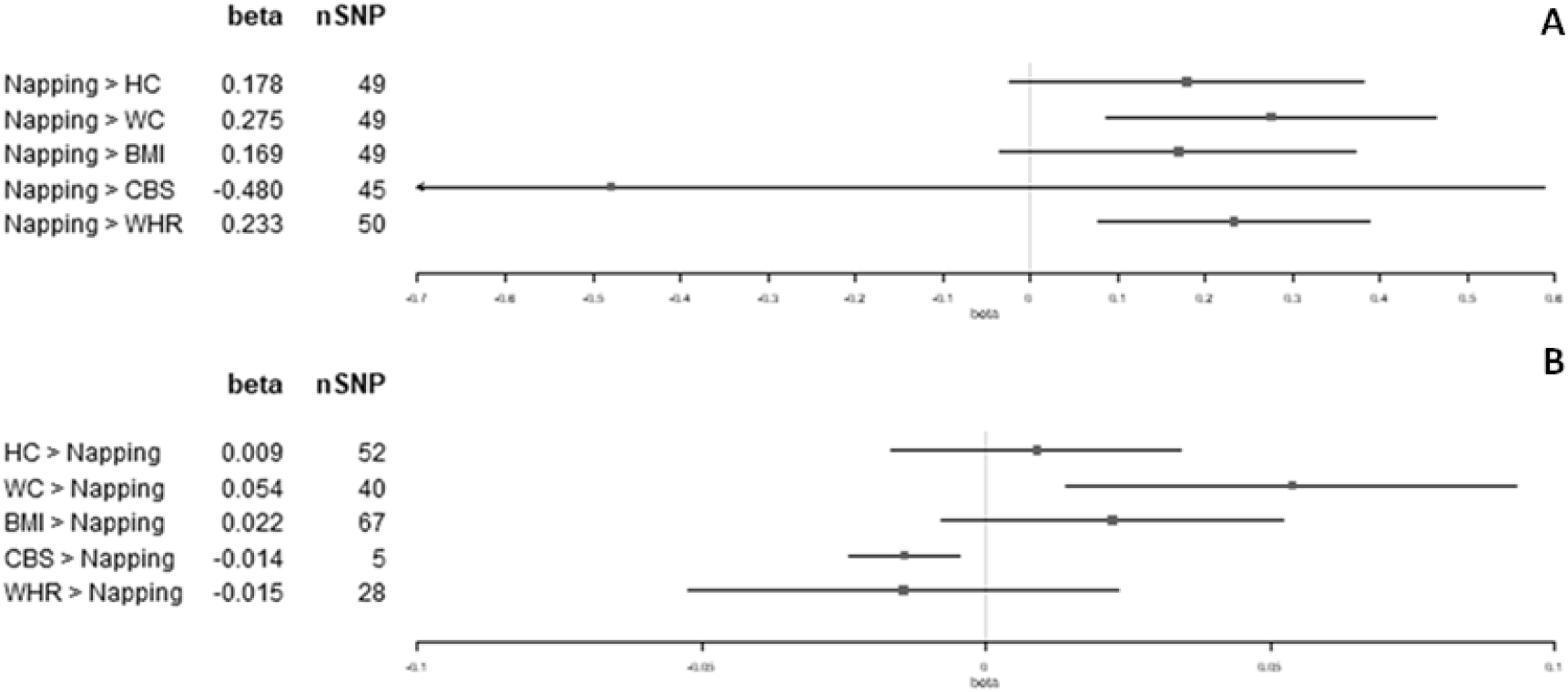
Two-sample bidirectional MR. Forest plot of A) napping trait effects on adiposity and B) Adiposity trait effects on napping. All results presented are IVW.

**Figure 5.**
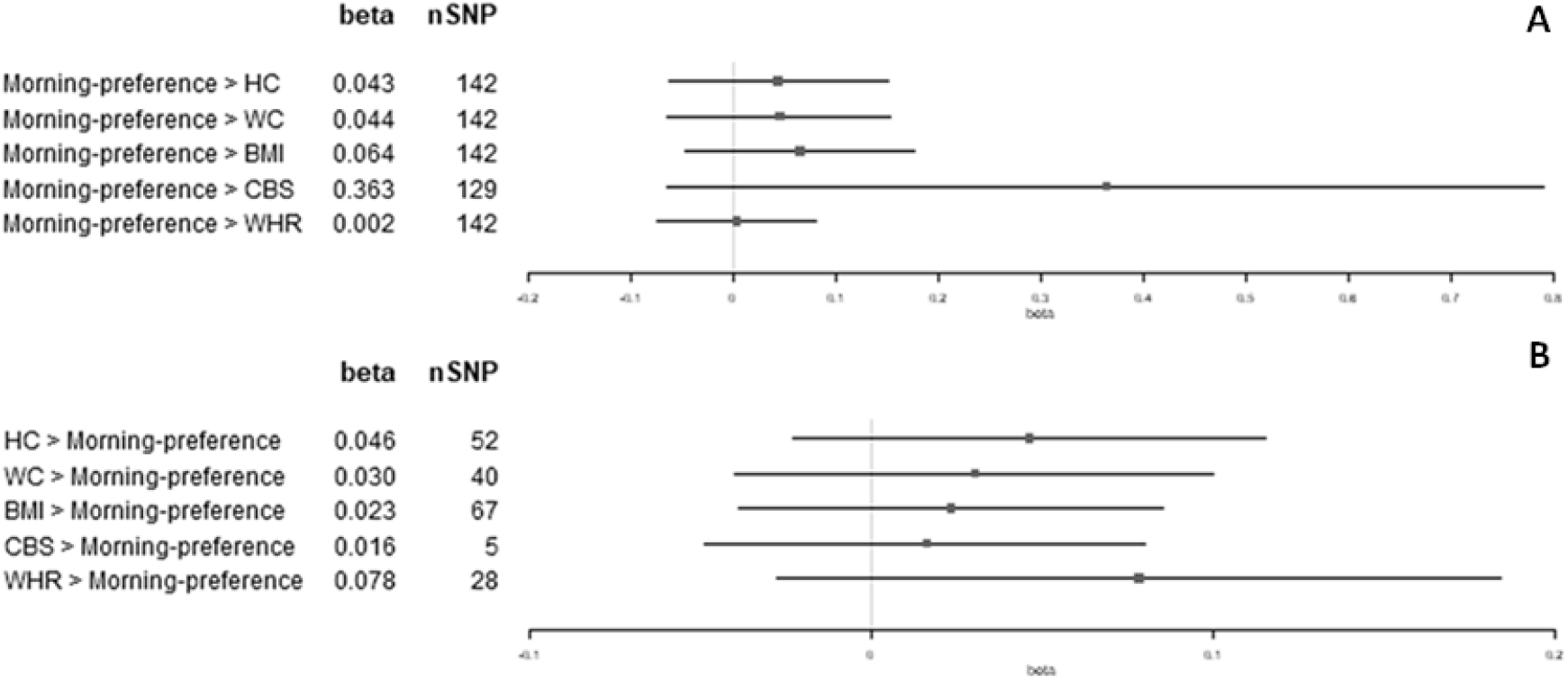
Two-sample bidirectional MR. Forest plot of A) napping trait effects on adiposity and B) Adiposity trait effects on napping. All results presented are IVW.

**Figure 6.**
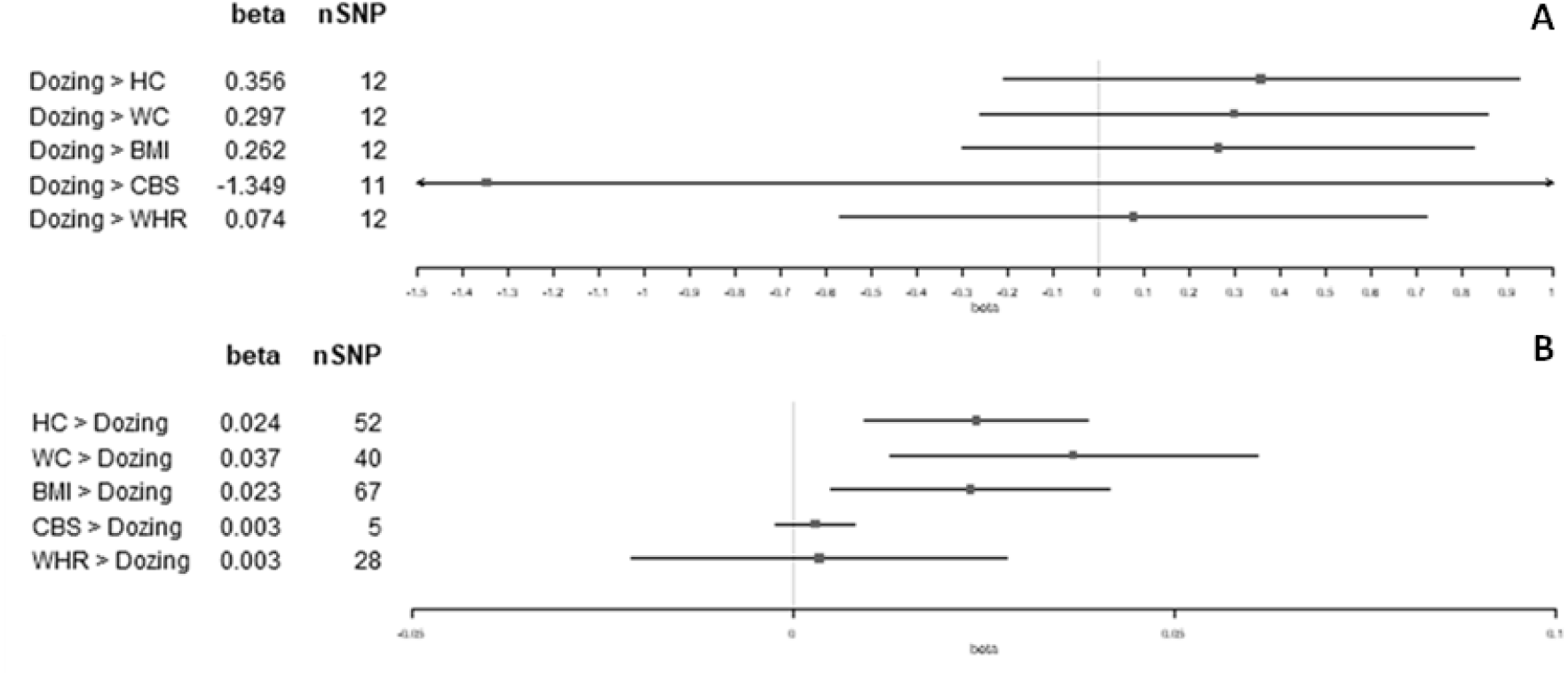
Two-sample bidirectional MR. Forest plot of A) daytime-sleepiness trait effects on adiposity and B) Adiposity trait effects on daytime-sleepiness. All results presented are IVW.

Results from most sensitivity analyses for effects of different sleep traits on adiposity were consistent with the main analysis results. With the exception of sleep duration effect on child-BMI, where the results were attenuated following Steiger-filtering (beta=-1.77 SD, 95% CI=-7.24, 3.39), although they remained directionally consistent with those reported in the main analysis (**Supp. Table 4**).

R^2^ values suggested that the instruments explained 0.13-2.07% of the variance of the exposures and genetic variants contributing to the sleep trait instruments had a combined F-statistic of 13.2-48.4, indicating reasonable instrument strength[29,40]. Between-IV heterogeneity for sleep trait instruments ranged from 0 – 88% (Qstat = 19-1126; Qpval = 3.6×10^−153^ – 4.8×10^−1^). Weighted and unweighted I^2^_GX_ [36] was also calculated for each of the sleep traits on adiposity traits and found to be between 0 – 74%. SIMEX corrections [37] were conducted for each analysis to account for this and found to be largely consistent with MR Egger results reported in the main analyses (**Supp. Table 3**).

Per SD increase in HC (1 SD = 9.2cm), we observed a category increase in daytime-sleepiness (beta=0.02 SD, 95% CI=0.01, 0.04) (**Fig 6**). Per SD increase in WC (1 SD = 12.5cm), we observed a category increase in both napping and daytime-sleepiness (beta=0.05 SD, 95% CI=0.01, 0.09 and beta=0.04 SD, 95% CI=0.01, 0.06 respectively) (**Figs 4 and 6**). Per SD increase in adult-BMI (1 SD = 5.1kg/m^2^), we observed a category increase in daytime-sleepiness (beta=0.02 SD, 95% CI=0.00, 0.04) (**Fig 6**). Per SD increase in child-BMI (1 SD = 4.7kg/m^2^), we observed a category decrease in napping (beta=-0.01 SD, 95% CI=-0.02, 0.00) (**Fig 4**). Per SD increase in WHR (1 SD = 0.07), we observed a category decrease in insomnia symptoms (beta=-0.04 SD, 95% CI=-0.07, -0.01) (**Fig 2**). There was no other evidence found for effects of adiposity traits on sleep (**Figs 2 – 6**). For all analyses, results from MR-Egger, weighted median and weighted mode methods were similar to IVW estimates (**Supp. Table 5**).

Results from most sensitivity analyses for effects of different adiposity traits on sleep were consistent with the main analysis results (**Supp. Table 6**).

Genetic variants contributing to the adiposity trait instruments had a combined F-statistic of 43.2-68.2, indicating good instrument strength[29,40], and r^2^ values suggest that instruments explain 0.98-2.00% of the variance of the exposures. Between-IV heterogeneity for adiposity trait instruments ranged from 0 – 97% (Qstat = 3-399; Qpval = 3.8×10^−49^ – 3.6×10^−1^). Weighted and unweighted I^2^_GX_ [36] was also calculated for each of the adiposity traits on sleep traits and found to be between 0 – 91%. SIMEX corrections [37] were conducted for each analysis to account for this and found to be largely consistent with MR Egger results reported in the main analyses (**Supp. Table 5**).

## Discussion

### Summary of main findings

This study assessed the direction of effect between a series of adiposity and sleep traits using two-sample MR analyses. Overall, we found consistent MR evidence, including from radial and Steiger filtering, of insomnia symptoms increasing mean WC, BMI and WHR, with little evidence for an effect in the opposing direction of adiposity on insomnia. There was evidence that napping increased mean WHR, but no effect was found in the other direction. Our results suggest higher mean child-BMI results in lower odds of napping, and that longer sleep duration may result in lower child-BMI, though for the latter Steiger-filtering, suggested the presence of shared causal variants more strongly associated with child-BMI, and there was no evidence for an effect in the opposing direction of adiposity on sleep duration.

A bidirectional adverse effect was found between napping and WC, which was consistent across radial-MR and Steiger-filtered results. We found little evidence for an effect of daytime-sleepiness on adiposity in our main results; however, evidence for an adverse effect of daytime-sleepiness on HC, WC and BMI was found in radial-MR (and on WC and BMI in Steiger-filtered results). Reciprocal adverse effects were found in the opposing direction for the effect of HC, WC, and BMI on daytime-sleepiness, which was consistent across radial-MR and Steiger-filtered results.

### Public health and clinical implications

The public health and clinical implications of these results are potentially far-reaching. Our results show that experiencing more frequent insomnia symptoms increases BMI (**Fig 2a**). Therefore, someone who suffers from insomnia may struggle to lose weight without first dealing with their insomnia.

Overall, better understanding the complex relationship between sleep and adiposity traits may help individuals who struggle to maintain healthy sleep or healthy weight, improve overall health, and consequently reduce the economic burden to our healthcare system.

### Comparison with previous literature

The effects of insomnia on adult-BMI (1 SD = 5.1kg/m^2^) and WHR (1 SD = 0.07) found in this study (beta=0.47 SD, 95% CI=0.22, 0.73 and beta=0.34 SD, 95% CI=0.16, 0.52 respectively) are consistent, with but less conservative, than previously reported two-sample MR findings by Xiuyan *et al* (beta=0.08 SD, 95% CI=0.06, 0.09 and beta=0.03 SD, 95% CI=0.02, 0.04 respectively)[41]. This may be attributed to the different GWAS used for their insomnia exposure (a meta GWAS of UKB and 23andme vs UKB-only) and also differences in underlying sample population demographics. Our study used only participants of European descent, whereas Xiuyun *et al* used a ‘mixed’ population for their WHR GWAS. Another study by Dashti *et al* also explored the effects of insomnia on BMI (1 SD = 5.1kg/m^2^), of which the results were consistent with those reported here (beta=0.36 SD, 95% CI=0.26, 0.46)[15].

The effect of sleep duration on child-BMI found in this study (beta=-0.93 SD, 95% CI=-1.74, -0.11) is consistent, but less conservative, than the previous reported two-sample MR findings (1 SD = 4.65 kg/m^2^) (beta=-0.27 SD, 95% CI=-0.51, -0.02)[17]. The same study by Wang *et al* also tested robustness of this effect in supplementary analyses by correction with MR-PRESSO and found consistent results (beta=-0.31, 95% CI=-0.53, -0.01), whereas our study reported some attenuation of effect following removal of SNPs more strongly associated with the outcome in Steiger-filtering (beta=-0.65 SD, 95% CI=-1.56, 0.25). We have also taken note of the imprecisely estimated MR-Egger, median and mode results, and the weighted and unweighted I^2^_gx_ result of 0% in our main analyses, suggesting a large amount of measurement error bias [36] (**Supp Table 3**).

The bidirectional effect between napping and WC found in this study (beta=0.28 SD, 95% CI=0.09, 0.46 vs beta=0.05 SD, 95% CI=0.01, 0.09) is consistent with the previously reported two-sample MR findings by Dashti *et al* (beta=0.28 SD, 95% CI=0.11, 0.45 vs beta=0.03 SD, 95% CI=0.01, 0.07)[16]. The unidirectional effect we found for napping on WHR (beta=0.23 SD, 95% CI=0.08, 0.39) is also consistent with the previously reported results (beta=0.19 SD, 95% CI=0.04, 0.33), as to be expected given the same underlying data used for these analyses (UKB and GIANT summary statistics)[16]. Furthermore, our additional sensitivity analyses with radial-MR and Steiger-filtering found these associations to be robust.

To our knowledge, the unidirectional effect of child-BMI on napping found in this study (beta=0.28 SD, 95% CI=0.09, 0.46) has not previously been reported. No SNPs were flagged for removal in either radial-MR or Steiger-filtering for this analysis, but weighted and unweighted I^2^_gx_ was found to be 0-14%, suggesting a large amount of measurement error bias [36] (**Supp Table 4**).

The unidirectional effect of adult-BMI on daytime-sleepiness reported in the main results of this study (beta=0.02 SD, 95% CI=0.00, 0.04) is consistent with that previously found by Dashti *et al* (beta=0.02 SD, 95% CI=0.01, 0.03)[15], furthermore this result persists after removal of outliers in radial-MR (beta=0.02 SD, 95% CI=0.01, 0.04). HC and WC were also found to increase daytime-sleepiness (beta=0.02 SD, 95% CI=0.01, 0.04 and beta=0.04 SD, 95% CI=0.01, 0.06 respectively), both of which were robust to radial-MR and Steiger-filtering analyses. Whilst our main results in the opposing direction report little effect of daytime-sleepiness on adult-BMI, HC or WC, this may be attributed to the moderate to high heterogeneity between SNPs for this instrument (I^2^ = 59-73%), leading to imprecise estimation. Following the removal of one outlier SNP (rs6741951) from the daytime-sleepiness instrument in radial-MR, heterogeneity between SNPs was reduced to 0-21%, and an adverse effect was then found for adult-BMI (beta=0.48 SD, 95% CI=0.13, 0.82), HC (beta=0.55 SD, 95% CI=0.12, 0.97) and WC (beta=0.51 SD, 95% CI=0.14, 0.88). Altogether, the evidence suggests that a bidirectional relationship may exist between daytime-sleepiness and adult-BMI, WC and HC.

### Strengths and limitations

A key strength of this study is the use of two-sample MR to systematically appraise the causal effects of each of our sleep traits on adiposity and vice versa. Furthermore, MR assumptions were thoroughly tested with the use of additional sensitivity analyses, such as radial-MR and Steiger-filtering, the results of which provide evidence for the robustness of our results. The genetic summary data used for all traits in this study were obtained from the largest available GWAS whilst still maintaining zero overlap between exposure and outcome datasets.

While we were thorough in our assessment of MR assumptions using various sensitivity analyses, we were not able to directly appraise independence of IVs from potential confounding factors. Given that MR uses germline IVs, it is largely understood that these will not be influenced by confounders. Minimising population stratification may help to alleviate concerns of independence assumption violation[42], but this is difficult to test in a two-sample MR framework.

The use of overlapping sample populations between exposures and outcomes in a two-sample MR setting may be a potential source of bias[43]. Therefore, despite the availability of GWAS with larger sample sizes generated from meta-analysis of UKB and either GIANT or EGG data, we opted to use GWAS that utilised UKB-only sample populations for our sleep traits and GIANT-only or EGG-only sample populations for our adiposity traits to ensure zero sample overlap between exposure and outcomes for our analyses.

### Further work

The analyses presented here demonstrate robust casual evidence for both uni- and bi-directional relationships between sleep and adiposity, therefore further investigation is required to inform clinical guidelines and policy. To improve the robustness of the findings in this study, it would be interesting to investigate the associations found using objective measures that correspond to self-report sleep traits, such as accelerometer-derived sleep duration vs self-report sleep duration[44]. Furthermore, genetic epidemiological studies are disproportionately conducted in population samples of European ancestry. Therefore, future studies that include populations from a variety of ancestries will only serve to better our understanding of the genetics that underpin these associations.

In this study, directionality was explored between sleep and adiposity. Moving forward, it would be interesting to use these results to inform and conduct mediation analyses to look at effects on outcomes such as cancers and cardiovascular disorders.

## Conclusion

This study has extended previous findings regarding the effect of sleep on adiposity and vice versa and provided robust evidence for these associations across a variety of methods. Collectively, the effect of insomnia on adiposity, and adiposity on daytime-sleepiness suggests that poor sleep and weight gain may contribute to a feedback loop that is detrimental to the overall health of the individual. Further understanding of these interactions and how, together, they might impact disease outcomes would be highly beneficial and informative for intervention studies seeking to improve overall health and consequently reduce the economic burden on our healthcare system.

## Data Availability

Summary data for the morning-preference, insomnia, sleep duration, napping, and daytime-sleepiness GWAS used in this study are available at:
https://sleep.hugeamp.org/dinspector.html?dataset=GWAS_UKBB_eu
Summary data for the adult-BMI, WC, HC and WHR GWAS used in this study are available at:
https://portals.broadinstitute.org/collaboration/giant/index.php/GIANT_consortium_data_files
Summary data for the child-BMI GWAS used in this study is available at:
http://egg-consortium.org/childhood-bmi.html

https://sleep.hugeamp.org/dinspector.html?dataset=GWAS_UKBB_eu

https://portals.broadinstitute.org/collaboration/giant/index.php/GIANT_consortium_data_files

http://egg-consortium.org/childhood-bmi.html

## DATA AVAILABILITY

This research was conducted using the UK Biobank Resource under application number 16391.

Summary data for the morning-preference, insomnia, sleep duration, napping, and daytime-sleepiness GWAS used in this study are available at: https://sleep.hugeamp.org/dinspector.html?dataset=GWAS_UKBB_eu

Summary data for the adult-BMI, WC, HC and WHR GWAS used in this study are available at: https://portals.broadinstitute.org/collaboration/giant/index.php/GIANT_consortium_data_files

Summary data for the child-BMI GWAS used in this study is available at: http://egg-consortium.org/childhood-bmi.html

## ACKNOWLEDGEMENTS

Genetic associations were contributed by the Genetic Investigation of Anthropometric Traits (GIANT), Early Growth Genetics (EGG), and UK Biobank consortium investigators.

## Notes

FUNDING BH is funded by an Above & Beyond breast cancer legacy grant from University Hospitals Bristol NHS Foundation Trust (www.aboveandbeyond.org.uk). MV is supported by the University of Bristol Alumni Fund (Professor Sir Eric Thomas Scholarship). TR is supported by a National Institute of Health Research Development and Skills Enhancement Award (NIHR302363). RCR is a de Pass Vice Chancellor’s Research Fellow. RMM is supported by a Cancer Research UK (C18281/A19169) programme grant (the Integrative Cancer Epidemiology Programme) (www.cancerresearchuk.org/funding-for-researchers). BH, TR, RMM, DAL and RCR work in the Medical Research Council Integrative Epidemiology Unit at the University of Bristol supported by the Medical Research Council (MC_UU_00011/1 and MC_UU_00011/6) (www.mrc.ukri.org) and the University of Bristol. RMM and DAL are also supported by the National Institute for Health Research (NIHR) Bristol Biomedical Research Centre, which is funded by the National Institute for Health Research (NIHR) (www.nihr.ac.uk) and is a partnership between University Hospitals Bristol and Weston NHS Foundation Trust and the University of Bristol. DAL is an NIHR Senior Investigator (NF-0616-10102). The funders had no role in study design, data collection and analysis, decision to publish, or preparation of the manuscript.

CONFLICTS OF INTEREST I have read the journal’s policy and the authors of this manuscript have the following competing interests: DAL has received support from Roche Diagnostics and Medtronic Ltd for research unrelated to that presented here. TR has received grants from Daiichi-Sankyo and Amgen to attend educational workshops. All other authors have no competing interests to declare.

### Competing Interest Statement

DAL has received support from Roche Diagnostics and Medtronic Ltd for research unrelated to that presented here. TR has received grants from Daiichi-Sankyo and Amgen to attend educational workshops. All other authors have no competing interests to declare.

### Funding Statement

BH is funded by an Above & Beyond breast cancer legacy grant from University Hospitals Bristol NHS Foundation Trust (www.aboveandbeyond.org.uk). MV is supported by the University of Bristol Alumni Fund (Professor Sir Eric Thomas Scholarship). TR is supported by a National Institute of Health Research Development and Skills Enhancement Award (NIHR302363). RCR is a de Pass Vice Chancellor's Research Fellow. RMM is supported by a Cancer Research UK (C18281/A19169) programme grant (the Integrative Cancer Epidemiology Programme) (www.cancerresearchuk.org/funding-for-researchers). BH, TR, RMM, DAL and RCR work in the Medical Research Council Integrative Epidemiology Unit at the University of Bristol supported by the Medical Research Council (MC_UU_00011/1 and MC_UU_00011/6) (www.mrc.ukri.org) and the University of Bristol. RMM and DAL are also supported by the National Institute for Health Research (NIHR) Bristol Biomedical Research Centre, which is funded by the National Institute for Health Research (NIHR) (www.nihr.ac.uk) and is a partnership between University Hospitals Bristol and Weston NHS Foundation Trust and the University of Bristol. DAL is an NIHR Senior Investigator (NF-0616-10102). The funders had no role in study design, data collection and analysis, decision to publish, or preparation of the manuscript.

### Author Declarations

This study used only openly available human data that were originally available at: https://sleep.hugeamp.org/dinspector.html?dataset=GWAS_UKBB_eu https://portals.broadinstitute.org/collaboration/giant/index.php/GIANT_consortium_data_files http://egg-consortium.org/childhood-bmi.html

