## Supplementary Methods: A for "Establishing causal relationships between sleep and adiposity traits using Mendelian randomisation"

## A)

**Sleep Trait Questionnaires in UK Biobank**

At baseline, participants completed a touchscreen questionnaire, which included questions about their sleep behaviours.

Chronotype (morning or evening preference) was assessed in the question “Do you consider yourself to be?” with one of six possible answers: “Definitely a ‘morning’ person,” “More a ‘morning’ than ‘evening’ person,” “More an ‘evening’ than a ‘morning’ person,” “Definitely an ‘evening’ person,” “Do not know,” or “Prefer not to answer”, from which we derived a five category variable for chronotype where “Definitely a ‘morning’ person” and “Definitely an ‘evening’ person” define either extreme.

Sleep duration was assessed by asking: “About how many hours sleep do you get in every 24 hours? (including naps).” The answer could only contain integer values.

To assess insomnia symptoms, participants were asked: “Do you have trouble falling asleep at night or do you wake up in the middle of the night?” where responses “Never/rarely”, “Sometimes” or “Usually” were coded as a three category variable.

Frequency of napping was assessed based on the question: “Do you have a nap during the day?”, with possible responses “Never/rarely”, “Sometimes” and “Usually” coded as a three category variable.

The frequency of daytime-sleepiness using the question: “How likely are you to doze off or fall asleep during the daytime when you don’t mean to? (e.g.: when working, reading or driving)”, with the answers coded as a four category variable “never”, “sometimes”, “often”, or “all of the time”.

For all UKB-derived variables, answers “Prefer not to answer” were coded as missin**g.**
